# Predicting Traffic Accident Injury Severity Using Ensemble Machine Learning Models: Incident Level and Generalized Insights via Explainable AI

**DOI:** 10.64898/2026.04.13.26350778

**Authors:** Eddie Zhang, Omer Mermer, Ibrahim Demir

## Abstract

Road traffic accidents represent a global public safety crisis, necessitating advanced computational tools for accurate injury severity prediction and effective decision support. This study evaluates high-performing ensemble machine learning models, including AdaBoost, XGBoost, LightGBM, HistGBRT, CatBoost, Gradient Boosting, NGBoost, and Random Forest, using a comprehensive National Highway Traffic Safety Administration (NHTSA) dataset from 2018 to 2022. While all models demonstrated exceptional predictive accuracy, with HistGBRT achieving the highest overall accuracy of 92.26%, a defining achievement of this work is the perfect classification (100% precision and recall) of fatal injuries across all ensemble architectures. To bridge the gap between predictive performance and actionable intelligence, this research integrates SHapley Additive exPlanations (SHAP) to provide both global insights into dataset-wide risk factors and local, instance-specific rationales for individual crash events. The global analysis identified ethnicity, airbag deployment, and harmful event type as primary drivers of injury severity, while local force and waterfall plots revealed the precise “push and pull” of variables for specific incidents. The results offer a robust, interpretable framework for stakeholders tasked with improving traffic safety and mitigating crash-related harm.

## 1. Introduction

Road traffic accidents (RTAs) remain one of the most pressing public safety concerns worldwide, representing a leading cause of fatalities and injuries across all age groups. According to the World Health Organization (WHO), approximately 1.3 million individuals die annually as a result of traffic crashes, with up to 50 million more sustaining injuries each year (WHO, 2018). These accidents impose substantial human suffering and generate over $400 billion in global economic losses. In the United States alone, the National Highway Traffic Safety Administration (NHTSA) reported more than 18,000 traffic-related deaths in the first half of 2024, underscoring the continued prevalence and severity of road accidents despite ongoing advancements in traffic laws and infrastructure development (NHTSA, 2024). As populations and urbanization increase globally, ensuring traffic safety has become an urgent policy and research priority.

Multiple interrelated factors influence the likelihood and severity of injury in traffic accidents, including human behavior, road and vehicle conditions, environmental influences, and socio-economic status. Prior studies have demonstrated that variables such as poor visibility, flooding, wet or icy road surfaces, and reckless driving substantially elevate hazardous road conditions and accident risks (Chen et al., 2016; Akhlaghi et al., 2023; Ehsani et al., 2023; Alabbad et al., 2024). Furthermore, Giron et al. (2025) highlights the influence of post-crash healthcare access, indicating that individuals from lower-income communities often experience worse outcomes due to delayed or inadequate medical attention. These findings suggest that to enhance safety outcomes, a comprehensive understanding of all factors contributing to injury severity is essential, along with the deployment of targeted preventive strategies (Marcillo et al., 2022).

Road networks, particularly in countries like the United States with over 4 million miles of roadways, serve as critical infrastructure supporting economic growth and mobility (Burghardt et al., 2022). However, this expansive network faces growing challenges due to increased traffic volume, inconsistent maintenance, and varying road quality. These challenges create hazardous conditions that can increase the probability and severity of crashes (Chinowsky et al., 2013). As dependence on road transportation continues to rise globally, ensuring the safety of these networks becomes even more critical.

Technology has played an increasingly important role in addressing these issues. Advances in sensors, vehicle systems, and roadway management technologies offer new opportunities to reduce crash frequency and mitigate injury severity (Schulze & Koßmann, 2010). Nonetheless, one major challenge persists: effectively analyzing the large, heterogeneous, and often incomplete traffic crash data. Inconsistencies in data quality, region-specific reporting formats, and limited availability of high-resolution datasets hinder the development of broadly applicable predictive models (Torbaghan et al., 2022; Demir et al., 2015).

Overcoming these barriers necessitates innovative strategies such as leveraging crowdsourced data from mobile devices to fill observation gaps (Sermet et al., 2020), establishing standardized benchmark datasets to ensure model reliability across disciplines (Ebert-Uphoff et al., 2017), and utilizing web-based geovisual analytics platforms to communicate complex findings to stakeholders (Xu et al., 2019a). Analyzing such complex data and drawing robust conclusions requires modern computational techniques capable of handling high-dimensional and unstructured inputs (Agliamzanov et al., 2020; Ramirez et al., 2022).

Machine learning (ML) and artificial intelligence (AI) techniques have emerged as promising tools in addressing these challenges, offering the ability to detect complex, nonlinear patterns and interactions within high-dimensional data (Obasi & Benson, 2023). ML-based injury severity prediction models have been applied in various contexts, from rickshaw crashes in Pakistan (Ijaz et al., 2021) to traffic crash in US (Mostafa et al., 2025), and bicycle accidents in Israel (Birfir et al., 2023). These models, including decision trees, support vector classifiers (SVC), and random forests (RF), have demonstrated superior performance over traditional parametric models such as logistic regression, particularly in their predictive accuracy and ability to capture variable interactions (Rezapour et al., 2020; Ardakani et al., 2023; Xu et al., 2019b).

Azhar et al. (2022) identified key factors influencing injury severity in heavy vehicle crashes, demonstrating that random forest models achieved better accuracy than classification and regression trees, and offered useful insights for road safety improvements, such as the impact of lighting conditions. Bokaba et al. (2022) evaluated models like KNN and SVC in Gauteng, South Africa, finding RF to be the most accurate (99%). Iranitalab and Khattak (2017) compared several statistical and ML models including multinomial logit, RF, SVM, and KNN, identifying KNN as the most accurate. Infante et al. (2022), using crash data from Portugal, noted that ML models generally match statistical ones in balanced datasets but may not outperform them with smaller or imbalanced data. Despite these advances, ML models are often perceived as “black boxes,” making their predictions opaque and difficult for stakeholders such as policymakers and emergency responders to interpret or trust (Krajewski et al., 2021; Belle & Papantonis, 2021; Bayar et al., 2009).

While statistical models often assume linear relationships between variables, which limits their capacity to model complex systems (Wang & Kim, 2019), ensemble learning techniques have shown improved accuracy and robustness (Cgengula et al., 2023; Boo & Choi, 2022). Zhu et al. (2024) proposed a data augmentation and ensemble learning framework that identified critical causes of accidents (e.g., lane changes, turns) from high-dimensional, small-sample data. They demonstrated that an ensemble of LightGBM, RF, CatBoost, and XGBoost achieved a precision of 0.85. Zhao et al. (2025) introduced an integrated Ensemble Learning-Logit Model for automated vehicle accidents, reporting 93% accuracy and identifying higher automation levels (SAE 3–5) as reducing injury severity.

Wei et al. (2024) found that stacking classifiers outperformed others in truck-related accident prediction tasks. Jamal et al. (2021) showed that XGBoost outperformed traditional models for crash injury prediction in Saudi Arabia, identifying critical factors such as road conditions and collision types. Moreover, ensemble models have demonstrated strong performance in specific contexts, including vulnerable road user crashes (Wu et al., 2021), freeway accidents (Yang et al., 2022), and elderly pedestrian crashes (Guo et al., 2021). Other studies, such as those by Islam et al. (2022), Ceven and Albayrak (2024), and Panicker and Ramadurai (2022), found random forest and conditional inference forests to be particularly effective in predicting accident severity under varying road and crash conditions.

Interpretability in ML is especially important in safety-critical applications. While gradient boosting algorithms such as XGBoost offer strong predictive performance, their lack of transparency limits their real-world applicability (Artega et al., 2020; Cicek et al., 2023). In contrast, interpretable models like linear regression or decision trees are more accessible but often less accurate or flexible in handling complex, nonlinear data (Alabbad et al., 2022). This trade-off between accuracy and interpretability is a central challenge in the development of reliable traffic injury prediction systems.

To address this challenge, explainable artificial intelligence (XAI) methods have become increasingly prominent. SHapley Additive exPlanations (SHAP), based on cooperative game theory, is one of the most widely used XAI techniques. SHAP attributes a unique contribution score to each input feature for every model prediction, enabling both global (dataset-wide) and local (instance-specific) interpretability (Lundberg & Lee, 2017; Saeed & Omlin, 2023). Numerous studies have successfully integrated SHAP with traffic injury severity models (Peng et al., 2024; Zahid et al., 2024; Rifat et al., 2024). For instance, Ahmed et al. (2023) used SHAP with ensemble models to reveal the importance of features such as vehicle count and road category in New Zealand data.

Kashifi (2024) applied XGBoost-SHAP to identify key risk factors in two-wheeler crashes, including helmet use and urbanization levels. Wu et al. (2016) and Hasan et al. (2024) highlighted how risk factors vary between rural and urban environments. Wei et al. (2023) used SHAP to explore the interactions between road geometry, speed distribution, and weather, finding those factors crucial for severe crashes. Cicek et al. (2023) applied SHAP to MLP models and identified alcohol use, speeding, and seatbelt noncompliance as major contributors. Amini et al. (2022) employed SHAP TreeExplainer to generate ranked feature importance lists across various car crash contexts.

While global SHAP analysis is valuable for identifying overall trends, local SHAP explanations, which focus on individual crash events, are applied far less frequently in traffic injury severity modeling. This gap limits the practical utility of ML predictions in operational settings, where first responders, law enforcement, and transportation planners require instance-level insights to make timely and informed decisions. Local SHAP visualizations, such as force plots, enable an understanding of how specific input features contribute to a single prediction, highlighting both the direction and magnitude of each variable’s effect. Studies such as Merabet et al. (2025a) emphasized the importance of local interpretability in domains characterized by high variability and complex interactions, such as solar radiation forecasting, where reliance on global trends can obscure critical case-specific nuances.

Similarly, Merabet et al. (2025b) applied local SHAP analysis in water quality prediction models to reveal the influence of environmental and meteorological parameters on water quality variables, demonstrating the utility of local explanations for site-specific environmental monitoring. In the context of injury severity prediction, Mermer et al. (2026) employed local SHAP analysis to uncover the influence of temporal, personal, and external factors on injury outcomes in agricultural accidents. Collectively, these findings support the broader argument that instance-level interpretability is essential for translating model outputs into context-aware, real-time decision-making tools.

In the context of traffic safety, the lack of local SHAP usage reduces the actionable value of ML outputs in high-stakes scenarios. Although ensemble models such as XGBoost, LightGBM, and CatBoost are widely adopted due to their superior accuracy, few studies have systematically compared these algorithms on real-world crash datasets using both predictive performance and interpretability as key evaluation metrics (Zhao et al., 2025; Wei et al., 2024). Without local interpretability, stakeholders are left with generalized findings that may not translate effectively to specific accidents or intervention strategies. Thus, incorporating both global and local SHAP analyses is essential for bridging the gap between model performance and meaningful, context-aware decision support.

To address these gaps, the present study implements and evaluates most widely used and high-performing ensemble machine learning models for predicting traffic accident injury severity using a real-world dataset provided by the National Highway Traffic Safety Administration (NHTSA). This dataset incorporates a wide range of driver, vehicle, environmental, and contextual features including time of day, vehicle/collision information, alcohol/drug levels and crash location. Unlike many previous studies that focus solely on predictive performance, this research integrates SHAP to achieve both global and local interpretability. This dual-level explainability enables not only the identification of key risk factors at a population level but also provides accident-specific rationales that can be used by emergency responders and policy analysts in real time.

The contributions of this study are threefold. First, it offers a comparative evaluation of several high-performing ensemble ML algorithms for injury severity prediction, highlighting their strengths and limitations. Second, it employs explainable AI techniques to generate comprehensive global and local feature importance explanations, bridging the gap between model performance and interpretability. Third, the study proposes a generalizable framework for the deployment of interpretable ML tools in safety-critical domains, supporting faster and more informed decision-making by stakeholders tasked with improving traffic safety and reducing crash-related harm.

The remainder of this paper is organized as follows: Section 2 presents the dataset, feature engineering process, machine learning models, and SHAP implementation. Section 3 outlines the experimental results including model performance, training efficiency, and explanation outcomes. Section 4 concludes with major findings, policy implications, and directions for future research.

## 2. Methodology

### 2.1. Data Description

The dataset used in this study was obtained from the National Highway Traffic Safety Administration (NHTSA), which provides publicly available records of vehicle-related accidents occurring in the United States. This dataset spans the years 2018 through 2022 and includes detailed information on accident circumstances, vehicle and roadway conditions, environmental influences, and personal attributes of those involved. The dataset incorporates both continuous and categorical variables. These variables encompass driver demographics (e.g., age, sex, seat position), accident context (e.g., time of day, location, weather), and crash mechanics (e.g., type of impact, use of restraints, airbag deployment). In total, the dataset comprises a rich set of variables that enables comprehensive modeling of accident injury severity. Table 1 summarizes the detailed descriptions of the cleaned and preprocessed dataset. It includes a mixture of continuous and categorical variables.

**Table 1.**
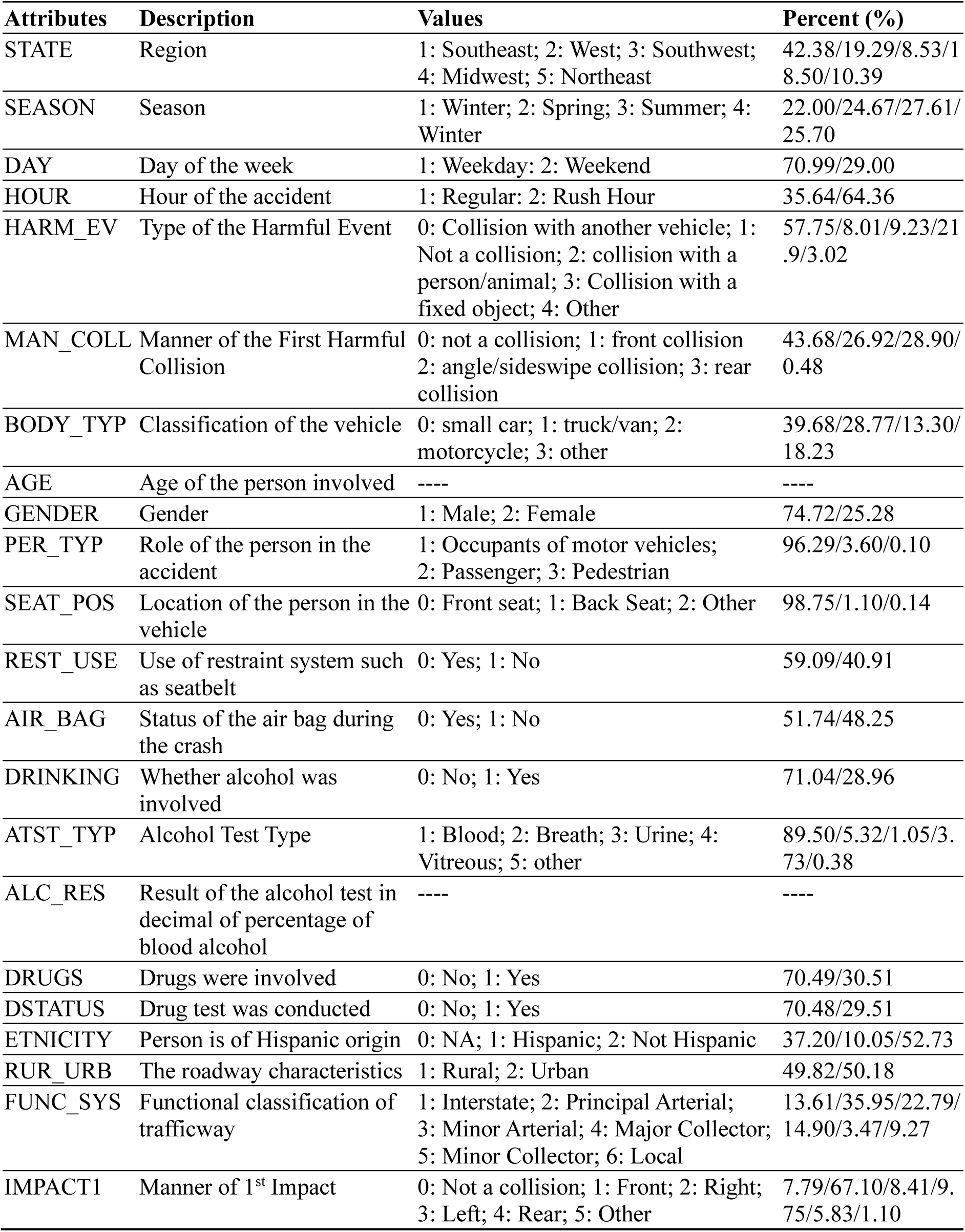
Detailed description of all variables in the dataset.

The categorical variables are represented as encoded integers and correspond to labels such as region, season, time of crash, collision type, vehicle type, and other driver- and environment-related features. For example, STATE represents one of five U.S. geographic regions (Southeast, West, Southwest, Midwest, and Northeast), while SEASON captures the season in which the crash occurred. Road conditions and the environment are reflected in variables such as RUR_URB (rural vs. urban setting) and FUNC_SYS (functional classification of the road). The AGE and ALC_RES (alcohol content in blood) variables are continuous, while attributes such as GENDER, DRINKING, and DRUGS are binary categorical variables. Usage of safety equipment is tracked through variables like REST_USE (restraint usage) and AIR_BAG deployment status. The inclusion of variables such as HARM_EV (first harmful event) and IMPACT1 (manner of first impact) allows for detailed modeling of crash dynamics.

In addition to the predictor variables, the target variable in this study is injury severity, which has been categorized into three levels: Property Damage Only (PDO), Injury, and Fatal Injury. These classes are derived from the original NHTSA classification schema, where PDO corresponds to “No Apparent Injury,” Injury includes cases such as “Possible Injury,” “Suspected Minor Injury,” and “Suspected Serious Injury,” and Fatal maps directly to “Fatal Injury.” This three-class schema allows for multi-class classification modeling and reflects real-world stratification in crash severity outcomes.

The frequency distribution of injury severity levels from 2018 to 2022, as detailed in Table 2, reveals significant fluctuations influenced by global events and changing traffic dynamics. A notable decline in total accident frequencies was observed during 2020 and 2021, coinciding with the COVID-19 pandemic and associated travel restrictions. However, a concerning post-pandemic trend emerged in 2022, characterized by a sharp increase in both the total number of accidents and the severity of outcomes. Specifically, the number of fatal injuries rose from 5,594 in 2018 to 7,916 in 2022, representing a substantial 41.5% increase over the five-year period. Throughout these years, the percentage of fatal injuries remained consistently high, accounting for 61% to 64% of the dataset’s annual records, which underscores the urgent need for the predictive and interpretable modeling framework developed in this study.

**Table 2:** Number of Fatal Injuries Per Year.

| Year | Injury Severity Category | Frequency | Percent (%) |
| --- | --- | --- | --- |
| 2018 | PDO | 1669 | 19% |
|  | Injury | 1630 | 18% |
|  | Fatal | 5594 | 63% |
| 2019 | PDO | 1925 | 18% |
|  | Injury | 2003 | 19% |
|  | Fatal | 6605 | 63% |
| 2020 | PDO | 1147 | 18% |
|  | Injury | 1307 | 21% |
|  | Fatal | 3918 | 61% |
| 2021 | PDO | 1091 | 18% |
|  | Injury | 1219 | 20% |
|  | Fatal | 3657 | 61% |
| 2022 | PDO | 2181 | 18% |
|  | Injury | 2236 | 18% |
|  | Fatal | 7916 | 64% |

These trends emphasize the importance of accurate and interpretable injury severity prediction. As such, this dataset serves as a robust foundation for developing ensemble machine learning models capable of capturing complex interactions among driver behavior, environmental context, and roadway characteristics. The preprocessing pipeline, including missing value removal, categorical encoding, outlier merging, and feature scaling, ensures that models are trained on a clean, balanced dataset that reflects the real-world conditions under which traffic accidents occur. This level of detail is critical not only for model performance but also for interpretability, especially when applying explainable AI methods such as SHAP to derive insights into both global feature importance and case-specific risk factors.

### 2.2. Machine Learning Models

#### Adaptive Boosting (AdaBoost)

AdaBoost, introduced by Freund and Schapire (1997), is the first and one of the most influential boosting ensemble methods. It operates on the principle that a strong classifier can be formed by sequentially combining multiple weak classifiers, each contributing to the final prediction. The algorithm employs a convex loss function and is known to be sensitive to noise and outliers in the dataset. AdaBoost adjusts the weights of training samples at each iteration, emphasizing misclassified instances to enhance the accuracy of subsequent weak classifiers (Hassan et al., 2017; Demir & Sahin, 2023). This iterative refinement continues until a specified classification error threshold is achieved, effectively reducing bias in complex prediction tasks when combined with other boosting strategies (Liu et al., 2015; Basaran et al., 2019).

Unlike some ensemble methods, AdaBoost is relatively resistant to overfitting and underfitting in certain classification contexts. It improves classifier performance by updating the sample distribution and iteratively incorporating only the most effective classifiers while discarding the weakest ones (Busari & Lim, 2021; Huang et al., 2021). This dynamic process leads to a cumulative increase in prediction accuracy. The detailed information and application of AdaBoost to investigate accident severity prediction can be found in various research articles, including AlMamlook et al., (2019). Dong et al., (2022), etc.

#### Extreme Gradient Boosting (XGBoost)

Extreme Gradient Boosting (XGBoost), introduced by Chen and Guestrin (2016), is a widely used ensemble learning algorithm based on decision trees, designed to enhance both computational efficiency and model flexibility in supervised learning tasks. XGBoost implements a sophisticated form of the gradient boosting framework by iteratively combining the predictions of multiple weak learners—typically decision trees—into a single, robust model. At each iteration, the model updates the previous learner by correcting its errors, using gradient-based optimization guided by a defined objective function to assess improvements in performance (Trizoglou et al., 2021). To reduce overfitting and improve generalization, XGBoost integrates regularization techniques that additionally support feature importance estimation, an essential factor for high-dimensional datasets (Jabeur et al., 2021).

The algorithm also employs a random sampling strategy to reduce variance and enhance the predictive power of the ensemble. Furthermore, XGBoost leverages both first- and second-order derivatives of the loss function, enabling more precise gradient direction estimation and thus more efficient minimization of the loss compared to traditional gradient boosting methods. Its support for parallel and distributed computing further contributes to reduced training times and more effective model exploration. XGBoost is highly efficient with large datasets and has been used in diverse fields from pedestrian injury severity prediction (Wu et al., 2024) to railroad accident analysis (Bridgelall & Tolliver, 2021).

#### Light Gradient Boosting Machine (LightGBM)

LightGBM is an advanced implementation of the gradient boosting decision tree (GBDT) framework that addresses several limitations traditionally associated with adaptive boosting methods, such as high computational complexity, limited memory efficiency, and long training times (Shehadeh et al., 2021). Built upon an ensemble of decision trees, LightGBM effectively handles both regression and classification tasks by leveraging the boosting paradigm (Chakraborty et al., 2020; Lu et al., 2023). Two notable techniques underpin its design: Exclusive Feature Bundling (EFB) and Gradient-based One-Side Sampling (GOSS), which jointly enhance efficiency and scalability (Ke et al., 2017).

To further reduce memory usage and training time, LightGBM employs a histogram-based algorithm along with a depth-constrained leaf-wise tree growth strategy. This approach improves prediction accuracy while mitigating overfitting risks associated with conventional GBDT models (Shehadeh et al., 2021). Additionally, LightGBM supports categorical features natively, avoiding the need for one-hot encoding, a requirement in many other gradient boosting frameworks (Ke et al., 2017; Lu et al., 2023). Recent studies have demonstrated the efficacy of LightGBM in road traffic injury severity, showing its applicability in real-world scenarios (Dong et al., 2022).

#### Histogram Gradient Boosting (HistGBRT)

Histogram-based Gradient Boosted Regression Trees (HistGBRT) is an advanced variant of the widely recognized gradient boosting ensemble method, commonly employed for both regression and classification tasks. Its primary goal is to enhance prediction accuracy by iteratively converting weak learners into a strong predictive model (Bentéjac et al., 2021). This is achieved through a sequential training approach, where each new weak learner is trained to correct the errors made by its predecessors (Sun et al., 2024). HistGBRT specifically addresses key limitations of traditional gradient boosting methods, such as prolonged training times on large datasets. This issue is mitigated by discretizing continuous input features into a finite set of values, which accelerates the training process (Marvin et al., 2023). Unlike other boosting techniques, HistGBRT stores continuous feature values in specialized containers and uses these to construct histograms during training, leading to improved efficiency and reduced memory consumption (Sun et al., 2024).

#### Categorical Boosting (CatBoost)

CatBoost, introduced by Dorogush et al. (2018), is a gradient boosting algorithm specifically designed to handle both numerical and categorical data efficiently. The name “CatBoost” is derived from “Category” and “Boosting,” reflecting its specialized capability in processing categorical features (Wu et al., 2019). Unlike traditional gradient boosting, CatBoost uses advanced algorithms to natively process categorical variables without extensive preprocessing, thus preserving the inherent structure of the data (Kulkarni, 2022). This makes it particularly effective when working with limited datasets or features dominated by categorical values.

CatBoost employs a unique tree structure known as an oblivious or symmetric tree, where each internal node at a given level splits the data using the same feature and threshold. This results in a tree of depth k having exactly 2^k^ leaves, enabling faster computations and easier interpretation (Wu et al., 2019; Lee et al., 2021). During training, CatBoost constructs a sequence of decision trees, each aimed at minimizing the loss function relative to its predecessor. To prevent overfitting, the algorithm includes parameter-based tree limits and early stopping, allowing training to stop if overfitting is detected before reaching the predefined number of iterations (Nguyen et al., 2022).

#### Gradient Boosting (GB)

The gradient boosting algorithm (GB) developed by Friedman (2001), is a decision-tree based ensemble learning technique that utilizes the boosting method to improve accuracy. Boosting works through training each model in an ensemble individually and iteratively, where each new model attempts to correct the errors made by previous models (Freund et al., 1999). Gradient boosting minimizes a loss function through the gradient descent method, where new trees or learners in a sequence are fitted to the residuals from the loss function with respect to the model’s current predictions (Natekin and Knoll, 2013). This allows the model to achieve increased predictive performance through focusing on the most difficult-to-learn data points (He et al., 2019).

Additionally, the algorithm also takes advantage of regularization using parameters such as learning rate and max_depth to help prevent overfitting through adding a penalty to a model’s complexity and improving robustness. These techniques result in a higher predictive performance for the gradient boosting algorithm, outperforming linear regression models and decision trees on non-linear, diverse, and complex datasets on both classification and regression tasks (Hosen and Amin, 2021).

#### Natural Gradient Boosting (NGBoost)

The Natural Gradient Boosting (NGBoost) algorithm is a probabilistic ML method that extends traditional gradient boosting by modeling predictive uncertainty in addition to generating point estimates (Duan et al., 2020). Unlike conventional boosting approaches, NGBoost predicts a full probability distribution for each data point, enabling estimation of outcome likelihoods (Zhou et al., 2024). While it follows the standard boosting framework of sequentially adding weak learners, NGBoost optimizes the negative log-likelihood of a chosen probability distribution rather than traditional loss functions such as mean squared error (Duan et al., 2020; Kavzoglu and Teke, 2021). NGBoost also employs natural gradients, which account the curvature of the loss function and enhance optimization stability (Martens, 2020). By updating distribution parameters at each iteration, NGBoost provides uncertainty estimates through the variance of the predicted distribution. This capability is especially valuable in high-risk applications such as medical and emergency decision making (Chen et al., 2022).

#### Random Forest (RF)

Random Forest (RF) is a widely adopted machine learning algorithm known for its strong performance in both classification and regression tasks (Breiman, 2001). Its effectiveness stems from the ensemble approach, where multiple randomized decision trees are constructed, and their outputs are aggregated, typically by averaging for regression or majority voting for classification. This methodology enhances predictive accuracy, particularly in high-dimensional settings where the number of variables exceeds the number of observations (Biau & Scornet, 2016). RF is also recognized for its flexibility, making it suitable for a wide array of learning tasks and scalable to large datasets. One of its key advantages is robustness to overfitting, achieved by averaging multiple de-correlated trees, which mitigates the risk of modeling noise from the training data (Breiman, 2001). Additionally, RF is resilient to outliers and can manage missing data without substantial performance loss. The algorithm provides interpretable metrics such as feature importance scores, which are valuable for understanding variable contributions. Recent advancements in crash severity prediction and related fields have widely used RF model, as shown by Zhang et al. (2018) and Zhang et al. (2022).

### 2.3. Model Development

The modeling process begins with acquiring and preparing the dataset obtained from the National Highway Traffic Safety Administration (NHTSA), encompassing a comprehensive set of accident-related variables. A rigorous data cleaning and preprocessing pipeline was employed to ensure high data quality and suitability for model development. This involved removing incomplete, inconsistent, and duplicate entries. Additionally, categorical values representing less than 0.1% of the dataset were grouped into a unified “Other” category to minimize data sparsity and noise. Categorical features were then encoded as integers ranging from 0 to n-1, where n denotes number of distinct categories per feature. Continuous variables were normalized using the min-max scaling to rescale values into the [0, 1] interval. This standardization, implemented via the MinMaxScaler, ensured equal features contribution and reduced potential boas arising from scale disparities (Mermer et al., 2025). Figure 1 presents the overall workflow implemented for machine learning (ML)-based traffic injury severity prediction.

**Figure 1:**
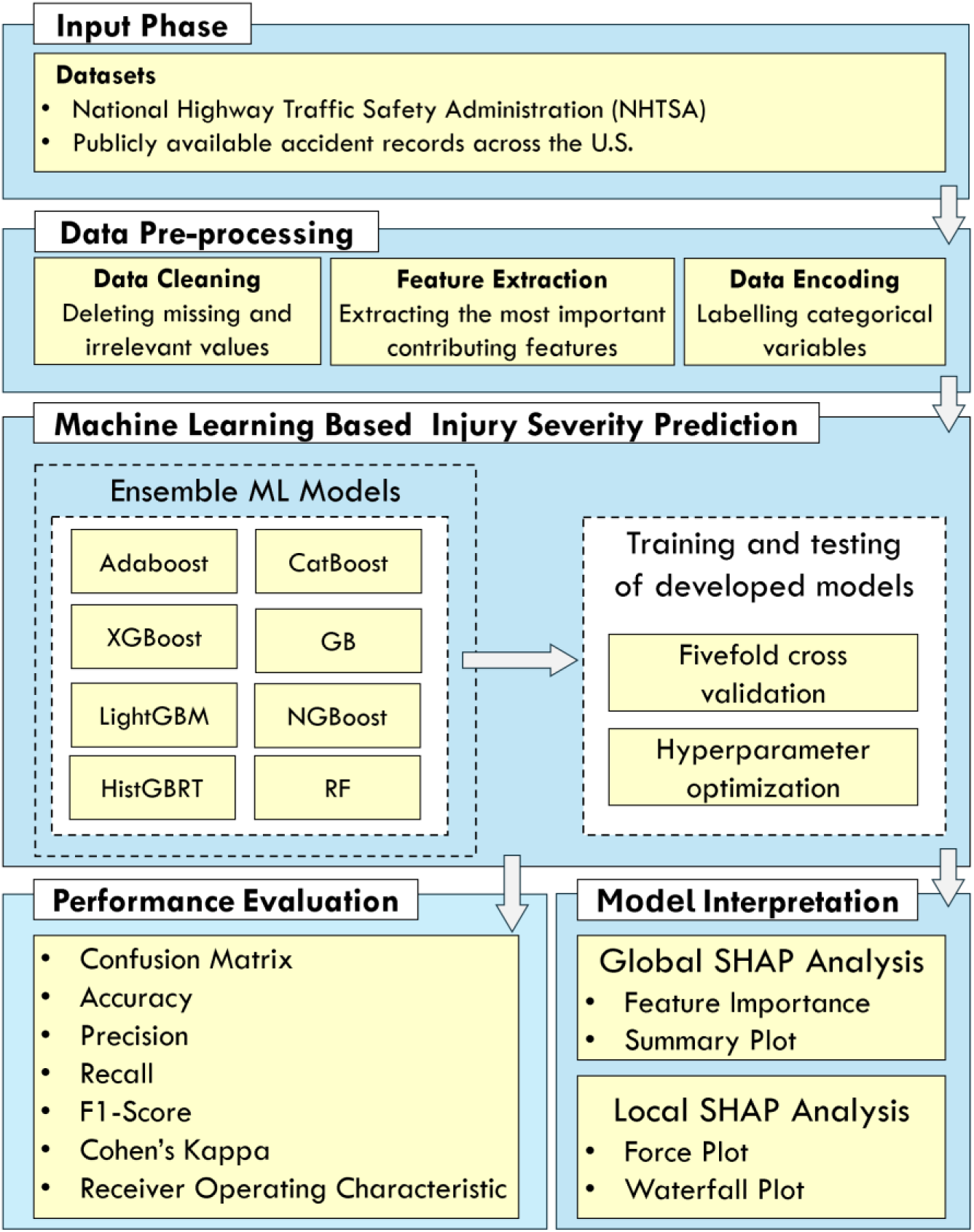
Architecture of the proposed framework

Following preprocessing, the cleaned dataset was randomly divided into a training set (80%) and a testing set (20%). This split allowed for robust model evaluation while preserving a sufficient number of samples for effective learning. Since the dataset comprises significantly more samples than features, dimensionality reduction was not aggressively pursued to retain critical information (Mermer et al., 2025). This decision aimed to maintain a broad representation of influencing factors while supporting the generalizability and robustness of the model.

Subsequently, a variety of ML models were selected and trained to predict injury severity levels. These models included tree-based ensemble algorithms that have shown superior performance in prior accident prediction studies. Hyperparameter optimization for each model was conducted using Bayesian Optimization, a probabilistic technique that efficiently explores the hyperparameter space. The optimization process was executed over 50 iterations for each model on the training set, and the best-performing configuration—based on validation accuracy—was retained for final testing. Table 3 provides a summary of the models and their optimized hyperparameters.

**Table 3.** Summary of training parameters of models developed in this study.

| **Models** | **Parameters** |
| --- | --- |
| Adaboost | Estimator = DecisionTree(Max_depth = 4, min_samples_split = 11, min_samples_leaf = 5, max_features = log2), n_estimators = 235, learning_rate = 0.0565 |
| XGBoost | N_estimators = 756, learning_Rate = 0.0866, max_depth = 4, min_child_weight = 6, gamma = 0.763, subsample = 0.866, colsample_bytree = 0.583, reg_alpha = $1.410 \times 10^{-6}$, reg_lambda = $1.609 \times 10^{-6}$, scale_pos_weight = 8.182 |
| LightGBM | N_estimators = 424, learning_rate = 0.0461, num_leaves = 210, max_depth = 3, min_child_samples = 57, subsample = 0.680, colsample_bytree = 0.677, reg_alpha = 0.00682, reg_lambda = 0.00209, boosting_type = “gbdt”, objective = “multiclass” |
| HistGBRT | Learning_rate = 0.0135, max_iter = 479, max_leaf_nodes = 235, max_depth = 4, min_samples_leaf = 29, l2_regularization = 0.0558, max_bins = 80, loss = “log_loss” |
| CatBoost | Iterations = 519, learning_rate = 0.0215, depth = 7, l2_leaf_reg = 0.00411, border_count = 188, random_strength = 1.10, bagging_temperature = 0.440 |
| GB | N_estimators = 201, learning_rate = 0.0593, max_depth = 4, min_samples_split = 3, min_samples_leaf = 10, subsample = 0.713, loss = “log_loss”, criterion = “squared_error” |
| NGBoost | N_estimators = 949, learning_rate = 0.0487, minibatch_frac = 0.543, col_sample = 0.731 |
| RF | N_estimators = 66, max_depth = 13, min_samples_split = 7, min_samples_leaf = 3, max_features = 0.183 |

### 2.4. Explainability Analysis with SHAP

As machine learning models grow in complexity and accuracy, the need for interpretability becomes increasingly critical, particularly in safety-sensitive domains such as traffic accident prediction. Among various explainable AI (XAI) techniques, SHAP (SHapley Additive exPlanations) has emerged as a state-of-the-art method for interpreting model outputs. Originally proposed by Lundberg and Lee (2017), SHAP is grounded in cooperative game theory and leverages Shapley values to attribute the contribution of each input feature to the model’s output. It provides a unified, model-agnostic framework capable of explaining predictions from any machine learning model, including ensemble and deep learning approaches (Aboulola et al., 2024; Yang et al., 2022; Demiray et al., 2025).

SHAP addresses a core challenge in machine learning by quantifying both the magnitude and direction of a feature’s influence on a specific prediction (Lundberg et al., 2020). This dual capability allows it to support both global and local interpretability. In global interpretability, SHAP provides summary visualizations that rank features by their overall importance across the entire dataset. These summary plots help identify the most influential variables and their relative contributions to prediction variance (Matrenin et al., 2024; Tiwari et al., 2024). In contrast, local interpretability involves examining individual predictions using force plots or decision plots, illustrating how each feature value contributed to a single prediction relative to a base value. This local perspective allows analysts and stakeholders to understand not only what the model predicted, but also why it made that specific prediction (Sahlaoui et al., 2021; Petrosian & Zhang, 2024).

SHAP is especially useful when applied to complex ensemble models like XGBoost, LightGBM, and CatBoost, which often yield high predictive performance at the expense of transparency. SHAP can reveal non-linear feature interactions, uncover hidden dependencies, and highlight variable thresholds that affect model outcomes, making it particularly valuable for traffic injury severity analysis where relationships between driver behavior, vehicle type, road conditions, and environmental factors are complex and multifaceted.

In this study, SHAP is used to evaluate feature importance and interaction effects in the ensemble models developed for predicting traffic injury severity. It enables both macro-level policy insights—such as identifying the most impactful safety variables—as well as micro-level operational guidance, such as understanding key risk factors for a specific crash event. This comprehensive explainability helps bridge the gap between predictive accuracy and actionable intelligence, offering practical value to first responders, transportation analysts, and policymakers. Its application in this study aims to improve both the diagnostic and prescriptive value of ensemble models by offering interpretable insights at both population and individual crash levels.

### 2.5. Model Evaluation

Evaluating model performance is essential to determining the predictive reliability and generalizability of any machine learning classifier. In this study, several standard evaluation metrics derived from the confusion matrix were employed to assess how well each model classified traffic injury severity cases. A confusion matrix is a square matrix in the R^m^ dimensional space, where m is the number of unique classes in the dependent variable. It provides a visual and quantitative summary of a model’s predictive performance by comparing actual labels to predicted ones during the testing phase. The confusion matrix consists of four fundamental elements: true positives (TP), true negatives (TN), false positives (FP), and false negatives (FN). These components serve as the foundation for computing several quantitative performance indicators.

▪ **TP (True Positive):** The number of instances where the predicted class matches the actual class (i.e., correctly predicted injury severity).

▪ **TN (True Negative):** The number of instances where all other class predictions are correctly identified as not belonging to the specific class.

▪ **FP (False Positive):** Instances where the model incorrectly assigns a positive label to a negative sample (i.e., wrong prediction of injury severity).

▪ **FN (False Negative):** Instances where actual positive cases are either missed or incorrectly classified by the model.

To ensure a comprehensive evaluation, this study computed the following metrics: accuracy, precision, recall (also referred to as sensitivity), and the F1-score. These metrics were selected for their ability to capture distinct aspects of classification performance, particularly in multi-class classification problems where class distribution may be imbalanced (Sokolova and Lapalme, 2009; Shreve et al., 2011). A summary of these evaluation metrics, along with their mathematical expressions and interpretative focus, is provided in Table 4.

**Table 4.**
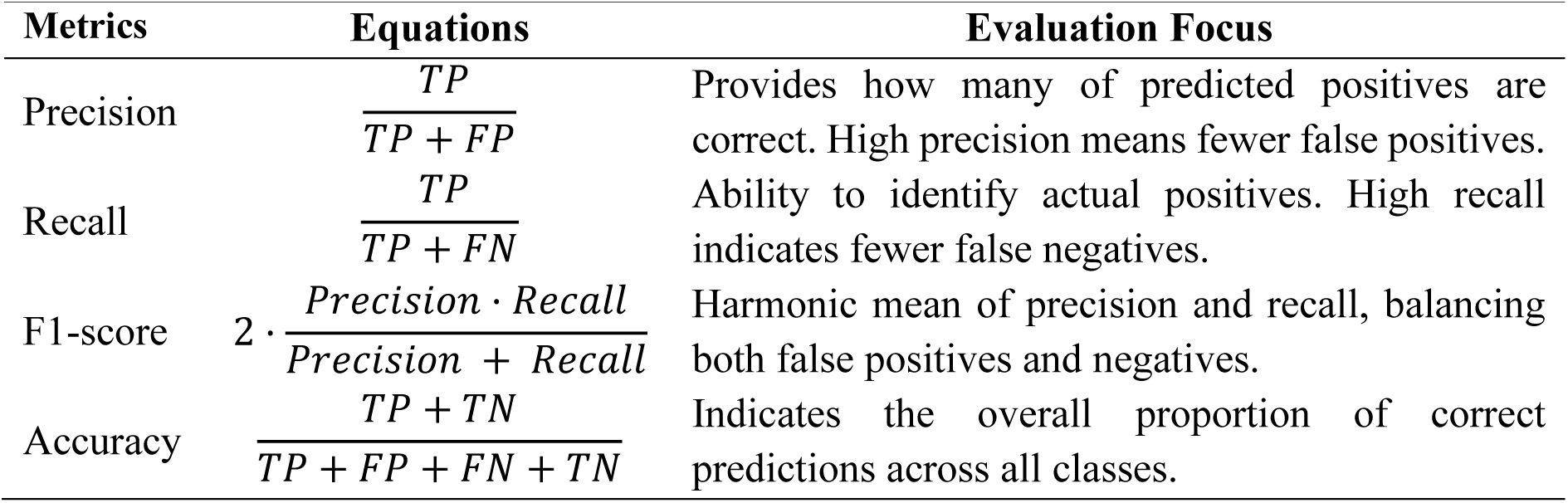
Summary of different performance evaluation metrics used.

In multi-class classification problems like injury severity prediction—where classes may include no injury, minor injury, and fatal injury—these metrics were calculated for each class individually and then aggregated using macro-averaging. This method assigns equal weight to all classes, thereby avoiding bias toward majority classes and enabling a fairer assessment across all injury levels.

Additionally, model performance was visualized using normalized confusion matrices, which highlight class-wise prediction strengths and misclassification patterns. These visualizations are especially useful in identifying weaknesses in predicting minority classes such as fatal injuries. A model might exhibit high overall accuracy while underperforming on critical but underrepresented classes, an important consideration in safety-critical domains like traffic crash analysis.

## 3. Results

The performance of the ensemble ML models in predicting agricultural injury severity was comprehensively evaluated using a combination of confusion matrices, quantitative metrics, ROC curves, and SHAP-based explainability analysis. The results indicate that while all models exhibit robust capabilities in classifying accident outcomes, they demonstrate unique strengths in handling the nuances of injury severity.

Table 5 presents the confusion matrices for each of the eight ensemble models, offering a detailed view of their classification outcomes for the three injury severity classes: Property Damage Only (PDO), Injury, and Fatal. In each matrix, the rows correspond to the actual number of accidents for a specific severity class, while the columns represent the predicted counts. Therefore, values along the main diagonal signify correct classifications, whereas off-diagonal values indicate misclassifications. The percentages in parentheses represent the class-specific recall, calculated as the proportion of correctly identified accidents out of the total actual accidents for that severity level.

**Table 5.** Confusion matrix generated for different ensemble ML models.

| Models | Actual Class | Predicted Class |  |  |  |  |  |
| --- | --- | --- | --- | --- | --- | --- | --- |
|  |  | PDO |  | Injury |  | Fatal |  |
| Adaboost | PDO | 1267 | (78.4%) | 349 | (21.6%) | 0 | (0%) |
|  | Injury | 335 | (20.06%) | 1335 | (79.94%) | 0 | (0%) |
|  | Fatal | 0 | (0%) | 0 | (0%) | 5533 | (100%) |
| XGBoost | PDO | 1267 | (78.4%) | 349 | (21.6%) | 0 | (0%) |
|  | Injury | 337 | (20.18%) | 1333 | (79.82%) | 0 | (0%) |
|  | Fatal | 0 | (0%) | 0 | (0%) | 5533 | (100%) |
| LightGBM | PDO | 1269 | (78.53%) | 347 | (21.47%) | 0 | (0%) |
|  | Injury | 351 | (21.02%) | 1319 | (78.98%) | 0 | (0%) |
|  | Fatal | 0 | (0%) | 0 | (0%) | 5533 | (100%) |
| HistGBRT | PDO | 1275 | (78.9%) | 341 | (21.1%) | 0 | (0%) |
|  | Injury | 342 | (20.48%) | 1328 | (79.52%) | 0 | (0%) |
|  | Fatal | 0 | (0%) | 0 | (0%) | 5533 | (100%) |
| Catboost | PDO | 1268 | (78.47%) | 348 | (21.53%) | 0 | (0%) |
|  | Injury | 337 | (20.18%) | 1333 | (79.82%) | 0 | (0%) |
|  | Fatal | 0 | (0%) | 0 | (0%) | 5533 | (100%) |
| GB | PDO | 1264 | (78.22%) | 352 | (21.78%) | 0 | (0%) |
|  | Injury | 349 | (20.9%) | 1321 | (79.1%) | 0 | (0%) |
|  | Fatal | 0 | (0%) | 0 | (0%) | 5533 | (100%) |
| NGBoost | PDO | 1261 | (78.03%) | 355 | (21.97%) | 0 | (0%) |
|  | Injury | 338 | (20.24%) | 1332 | (79.76%) | 0 | (0%) |
|  | Fatal | 0 | (0%) | 0 | (0%) | 5533 | (100%) |
| RF | PDO | 1265 | (78.28%) | 351 | (21.72%) | 0 | (0%) |
|  | Injury | 337 | (20.18%) | 1333 | (79.82%) | 0 | (0%) |
|  | Fatal | 0 | (0%) | 0 | (0%) | 5533 | (100%) |

A consistent and notable trend was observed across all models: a perfect classification rate for fatal cases. For every model, all 5,533 fatal injuries in the test set were correctly predicted, demonstrating an exceptional ability to identify the most severe outcomes. For the non-fatal classes, performance was also strong, though with some misclassification between them. For instance, the XGBoost model correctly predicted 1,267 PDO cases (78.4%) and 1,333 Injury cases (79.8%) but misclassified 349 PDO cases as Injury and 337 Injury cases as PDO. This pattern of confusion between the two less severe classes was similar across all models, with correct classification rates for PDO and Injury classes consistently hovering around 78-80%. Importantly, no PDO or Injury cases were incorrectly classified as Fatal, highlighting the models’ reliability in not overestimating severity.

While it is important to achieve an acceptable overall model performance, knowing the predictive performance by each injury severity class is fundamental for prioritizing and selecting appropriate treatment and mitigation measures. Table 6 provides a more detailed comparative analysis by injury severity class for all models. The perfect prediction for the fatal injury class is reflected in the 100% precision, recall, and F1-score achieved by all eight models for this category. Overall accuracy was remarkably high and consistent across the board, with most models achieving approximately 92.2%. The HistGBRT model registered the highest accuracy at 92.26%, while the GB model had the lowest at 92.05%, indicating only marginal differences in overall predictive power. The F1-scores for the PDO and Injury classes were also very stable, ranging from 78.29% to 78.87% for PDO and 79.03% to 79.61% for Injury. This demonstrates a well-balanced performance between precision and recall for the non-fatal classes, underscoring the robustness of the ensemble approaches for this classification task.

**Table 6.** Comparison of performance metrics by individual severity class.

| Models | Actual Class | Predicted Class |  |  |  |
| --- | --- | --- | --- | --- | --- |
|  |  | Precision (%) | Recall (%) | F1 Score (%) | Accuracy (%) |
| Adaboost | PDO | 79.09 | 78.40 | 78.74 | 92.24 |
|  | Injury | 79.28 | 79.94 | 79.61 |  |
|  | Fatal | 100.00 | 100.00 | 100.00 |  |
| XGBoost | PDO | 78.99 | 78.40 | 78.70 | 92.22 |
|  | Injury | 79.25 | 79.82 | 79.53 |  |
|  | Fatal | 100.00 | 100.00 | 100.00 |  |
| LightGBM | PDO | 78.33 | 78.53 | 78.43 | 92.09 |
|  | Injury | 79.17 | 78.98 | 79.08 |  |
|  | Fatal | 100.00 | 100.00 | 100.00 |  |
| HistGBRT | PDO | 78.85 | 78.90 | 78.87 | 92.26 |
|  | Injury | 79.57 | 79.52 | 79.54 |  |
|  | Fatal | 100.00 | 100.00 | 100.00 |  |
| Catboost | PDO | 79.00 | 78.47 | 78.73 | 92.23 |
|  | Injury | 79.30 | 79.82 | 79.56 |  |
|  | Fatal | 100.00 | 100.00 | 100.00 |  |
| GB | PDO | 78.36 | 78.22 | 78.29 | 92.05 |
|  | Injury | 78.96 | 79.10 | 79.03 |  |
|  | Fatal | 100.00 | 100.00 | 100.00 |  |
| NGBoost | PDO | 78.86 | 78.03 | 78.44 | 92.14 |
|  | Injury | 78.96 | 79.76 | 79.36 |  |
|  | Fatal | 100.00 | 100.00 | 100.00 |  |
| RF | PDO | 78.96 | 78.28 | 78.62 | 92.20 |
|  | Injury | 79.16 | 79.82 | 79.49 |  |
|  | Fatal | 100.00 | 100.00 | 100.00 |  |

The collective predictive performance of the eight ensemble classification algorithms is comparatively analyzed in Figure 2 using two critical metrics: Total Accuracy and Cohen’s Kappa. Total accuracy serves as a primary indicator of model success, representing the ratio of correctly predicted instances to the total number of cases. According to the study, the models exhibit remarkable consistency, with accuracy values clustering near 92.2%. The HistGBRT model achieves the highest precision at 92.26%, while the GB model registers the lowest at 92.05%, indicating only marginal variance in raw predictive power among the various architectures. Literature suggests that total accuracy provides a more robust measure of model prediction compared to simple average accuracy, particularly in multi-class scenarios.

**Figure 2:**
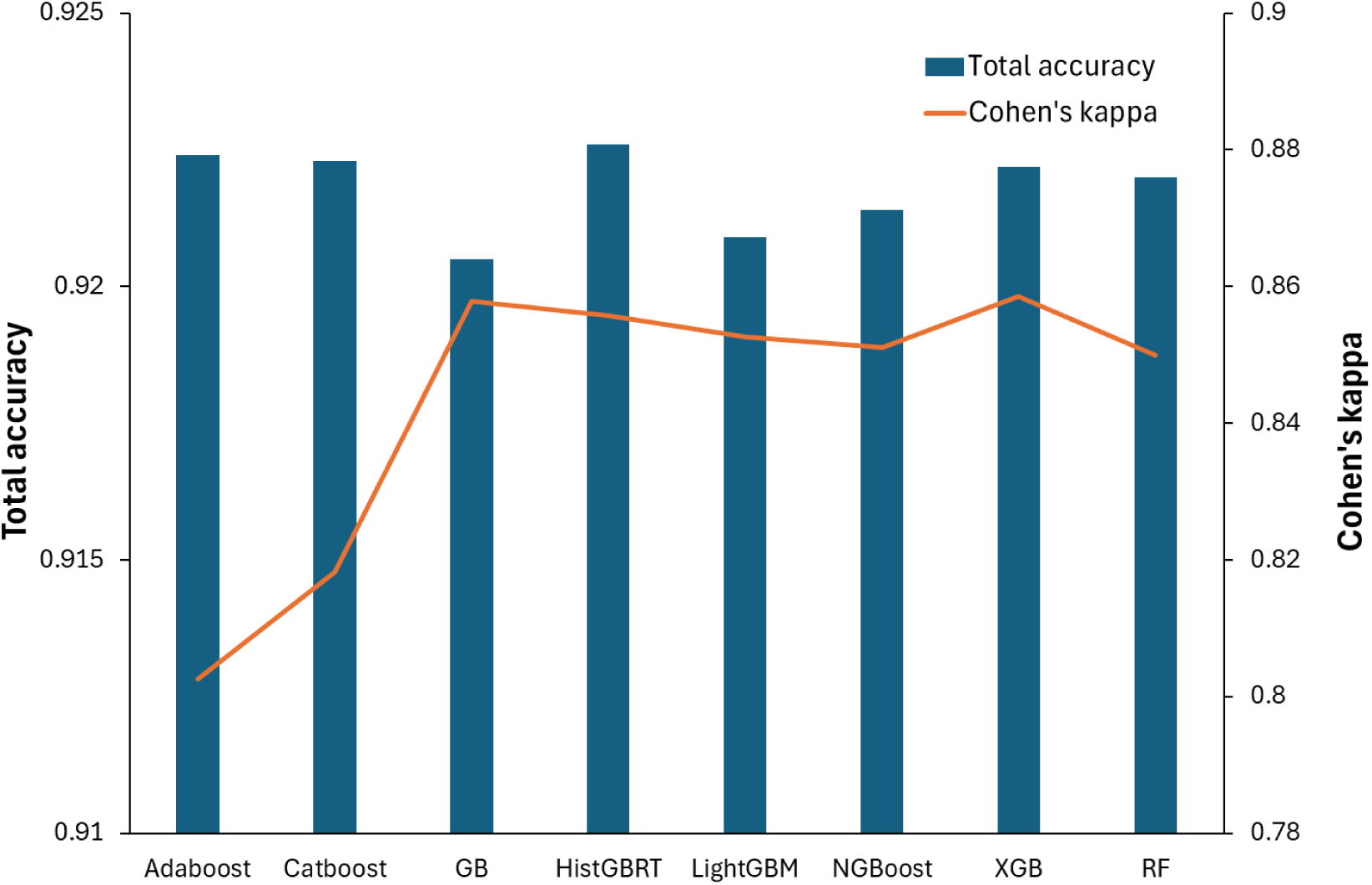
Models performance comparison based on overall accuracy and Cohen’s Kappa.

To further validate the reliability of these predictions, Cohen’s Kappa is utilized to measure the level of agreement between the actual injury categories and the model’s predictions. This statistic is essential for classification problems as it accounts for the possibility of agreement occurring by chance, with values ranging from 0 (random agreement) to 1 (complete agreement). As illustrated by the orange trend line in Figure 2, the Kappa values reflect “almost perfect” agreement, generally staying within the 0.80 to 0.86 range. Interestingly, while accuracy remains relatively flat across models, the Kappa scores show more distinct fluctuations. For example, XGBoost and GB demonstrate higher statistical agreement relative to their accuracy, whereas AdaBoost shows the lowest Kappa value despite high total accuracy. This comprehensive evaluation ensures that the models are not only achieving high hit rates but are also providing statistically sound classifications across the mutually exclusive categories of PDO, Injury, and Fatal injuries.

The discriminative performance of the eight ensemble models is visually and quantitatively confirmed through the Receiver Operating Characteristic (ROC) curves presented in Figure 3. Each model demonstrates superior classification ability, with curves positioned sharply in the upper-left quadrant, indicating a high true positive rate and a correspondingly low false positive rate across all classes. The Area Under the Curve (AUC) values provide a definitive measure of this performance; notably, every model achieved a perfect AUC of 1.00 for the fatal injury class (Class 2), representing flawless discrimination between fatal and non-fatal outcomes. For the non-fatal categories, the models maintained excellent precision, with AUC values approximately reaching 0.96 for PDO (Class 0) and 0.97 for Injury (Class 1) cases. These results significantly outperform a random-guess baseline (AUC = 0.5), validating that each tested algorithm effectively utilized the input features to differentiate between varying degrees of injury severity.

**Figure 3:**
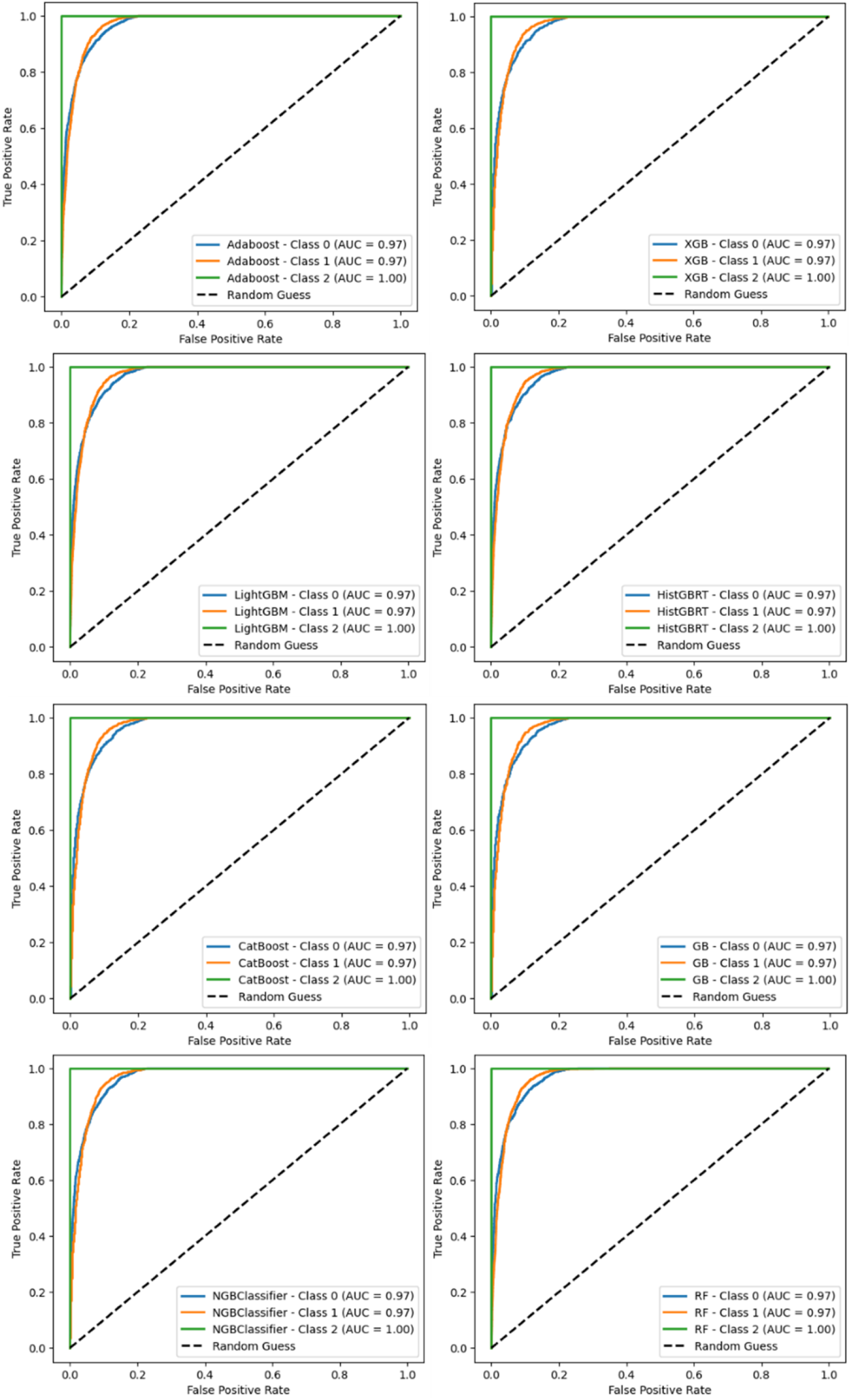
ROC curves of all models based on respective AUC scores for each class.

The SHAP interpretability analysis was performed at both global and local levels. The global interpretability analysis provided in Figure 4 identifies the primary drivers behind the models’ predictions by ranking features according to their mean absolute SHAP values. Across all ensemble architectures, Ethnicity consistently emerged as the most influential variable, often followed by Airbag deployment and the First Harmful Event (HARM_EV). For example, in the XGBoost and LightGBM models, Ethnicity held a mean SHAP value of approximately 0.59 to 0.60, significantly outweighing secondary factors like Restraint Use (REST_USE) and Manner of Collision (MAN_COLL). These results suggest that specific demographic and mechanical factors play a disproportionate role in determining injury severity outcomes within this dataset. By quantifying the magnitude of these contributions, the framework allows researchers to identify high-impact variables that are critical for population-level safety interventions.

**Figure 4:**
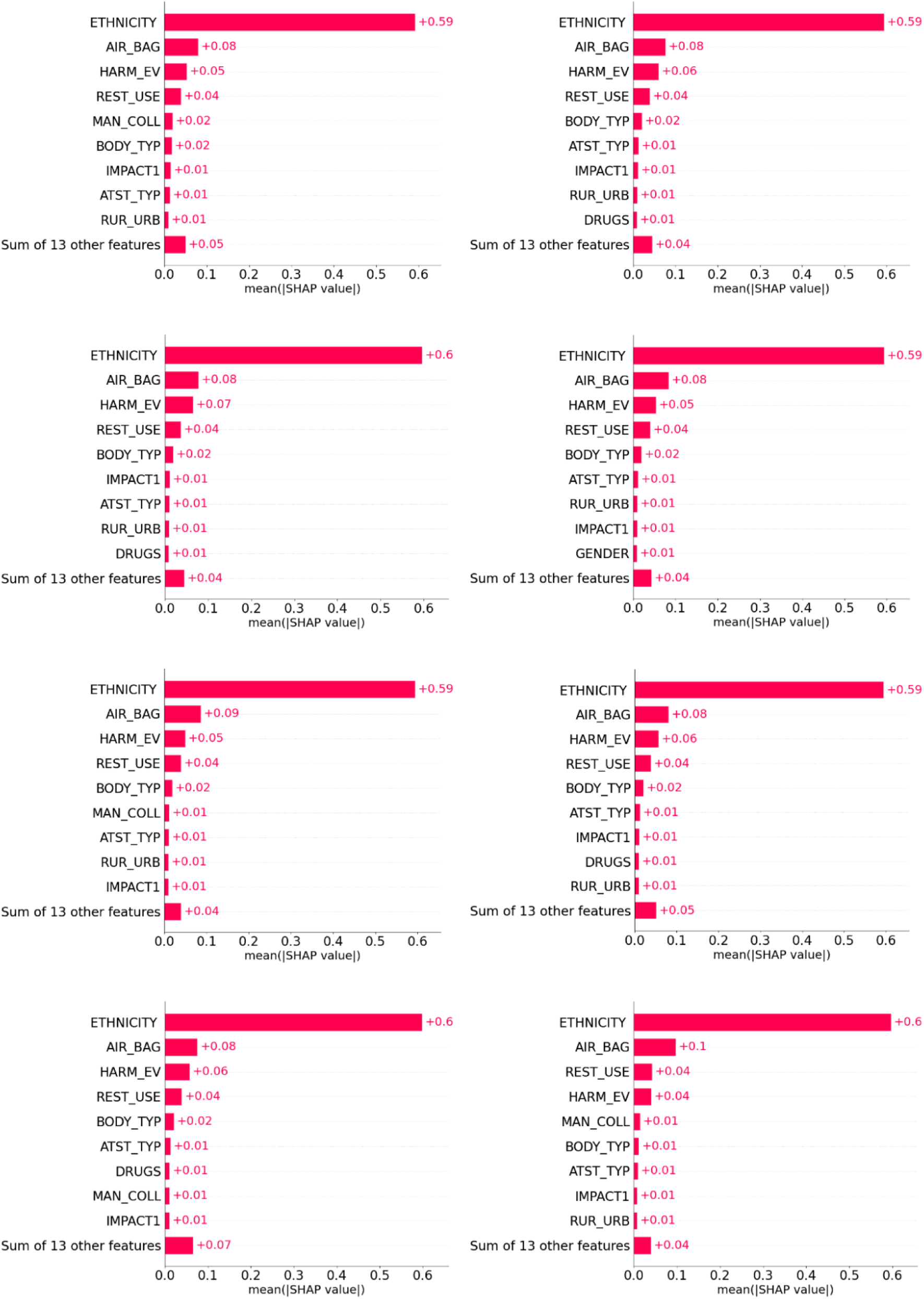
SHAP (bar plot) global interpretation for all models based on feature importance

While Figure 4 ranks importance, the SHAP summary plots in Figure 5 elucidate the complex relationship between feature values and the direction of their impact on model output. Each dot represents a single accident, where the color indicates the feature value (red for high and blue for low) and the x-axis position indicates the magnitude and direction of its impact. A distinct pattern is visible for the Ethnicity variable, where high values (represented in red) cluster on the positive side of the x-axis, indicating that these specific values push the model towards a higher severity prediction. Conversely, variables such as Airbag deployment and Restraint Use show a wider distribution, illustrating how their presence or absence can either mitigate or exacerbate the predicted injury level. These summary plots reveal non-linear feature interactions and hidden dependencies that traditional statistical models often fail to capture, providing policymakers with a nuanced view of how various risk factors, such as roadway characteristics and driver behavior, interact to influence crash outcomes.

**Figure 5:**
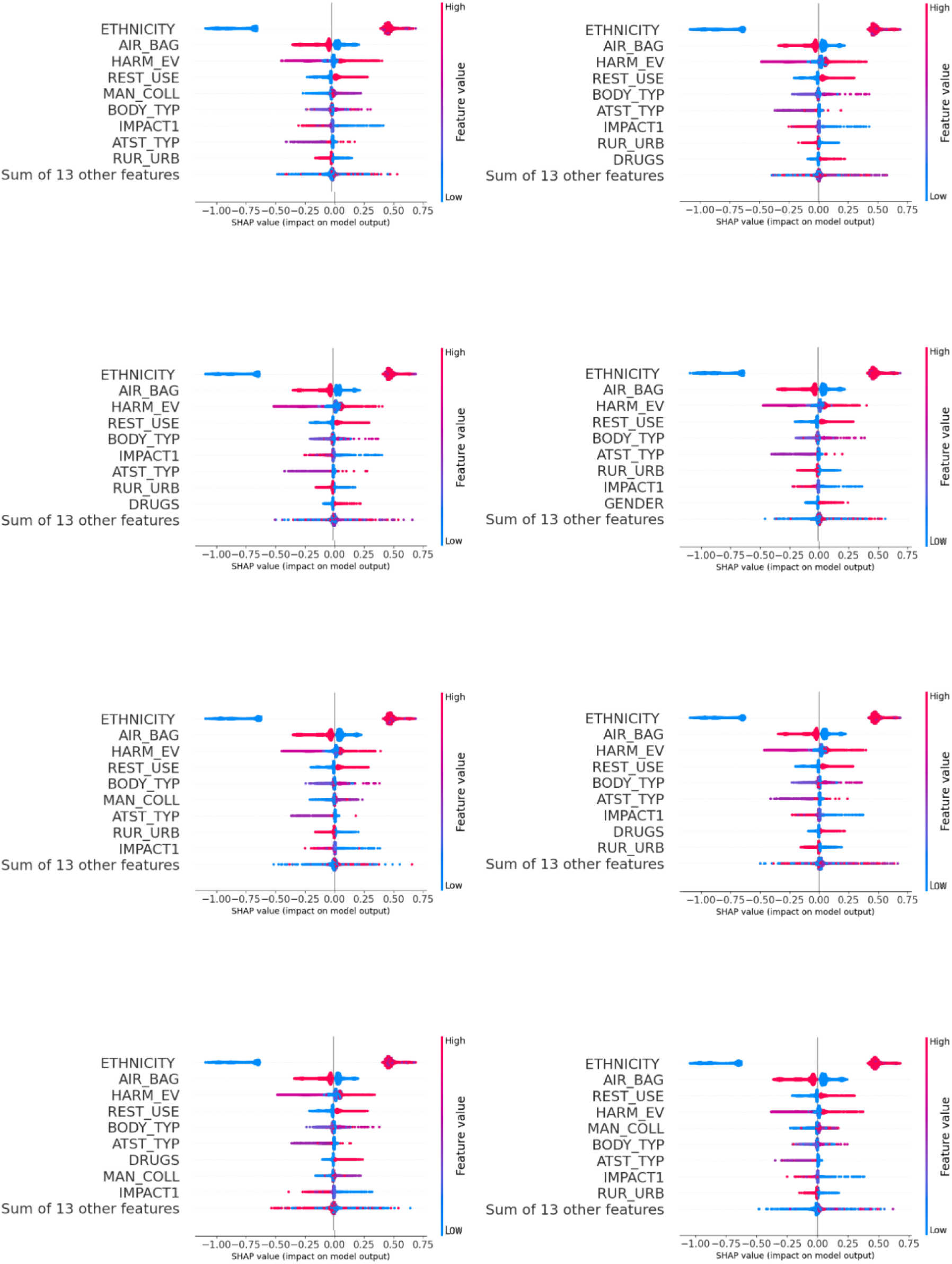
SHAP global interpretation for all models based on summary plot

The transition from population-level trends to accident-specific insights is achieved through the local interpretability analysis. Figure 6 shows the local interpretation based on force plot of chosen sample of XGBoost for predicting injury severity. Using force plots (right panel) and waterfall plots (left panel), the study provides a detailed breakdown of how individual features contributed to the prediction of a single crash event. The force plot in Figure 6 stresses the input variables for predicting injury severity and forcing accomplishment of XGBoost. Red color explains that particular input variables include more predictive control, whereas blue color supplies that the specific input variables involve lower predictive control. In samples such as Sample 5000 through 7000 (Figures 6f–h), the force plots show how the combination of high-impact variables, like Ethnicity, Restraint Use, and Impact Type, interact to “push” the final prediction toward a higher severity score. The length of the arrows in these plots corresponds to the magnitude of the SHAP value, offering a clear visual representation of feature strength for that unique accident.

**Figure 6:**
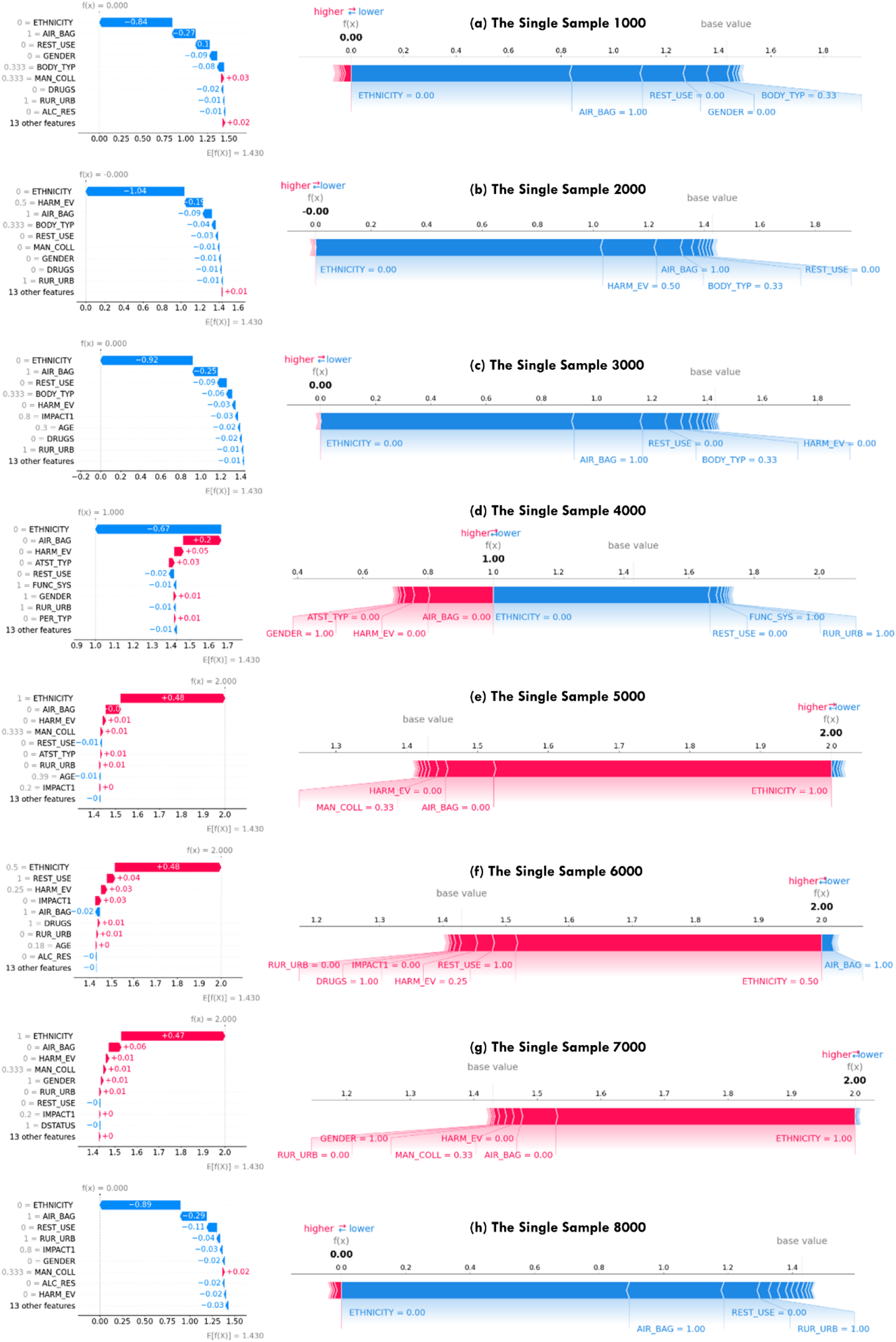
SHAP local interpretability for HistGBRT model and single samples prediction: force plot right panel and waterfall plots left panel.

The waterfall plots illustrate the path to a prediction, showing how each feature value for a specific sample, such as a driver’s age or a specific collision type, either increases or decreases the probability of a certain severity level relative to the base value. For example, in Sample 4000 (Figure 6e), the model visualizes how factors such as Ethnicity, Airbag Status, and Harmful Event type pull the prediction away from the baseline toward a specific severity level. Positive contributions (increasing severity probability) are typically indicated in red, while negative contributions (decreasing the probability) are shown in blue, allowing analysts to see exactly which variables were the primary drivers for that specific case.

By examining diverse samples (from Sample 1000 to 8000), Figure 6 demonstrates that the model’s reasoning varies significantly depending on the accident context. For instance, while one crash’s severity might be driven primarily by vehicle mechanics like airbag deployment, another might be influenced more heavily by driver-specific factors or roadway characteristics like RUR_URB (rural vs. urban). This level of detail provides context-aware rationales that are essential for safety-critical domains, enabling stakeholders, such as emergency responders and transportation planners, to identify specific risk factors in real-time and develop targeted intervention strategies based on the unique characteristics of an individual accident.

## 4. Discussion

The comprehensive evaluation of eight ensemble machine learning models in this study underscores their superior capability in predicting traffic accident injury severity, specifically within the context of rising fatal accidents observed between 2018 and 2022. The analysis revealed that while all models achieved high predictive accuracy, HistGBRT algorithm marginally outperformed others with an overall accuracy of 92.26%. However, the most significant finding was the ensemble ability of all eight models, including XGBoost, CatBoost, and RF, to achieve a perfect 100% recall and precision for fatal injuries.

This flawless discrimination of the most severe class, confirmed by an Area Under the Curve (AUC) of 1.00, represents a substantial advancement over traditional parametric models like logistic regression, which often struggle with the non-linear complexities and class imbalances inherent in crash data. The robustness of these findings is further validated by Cohen’s Kappa statistics ranging from 0.80 to 0.86, indicating that the high accuracy is not merely an artifact of chance but reflects a strong statistical agreement between the models’ predictions and actual injury outcomes. These results align with and elevate recent literature, such as findings by Zhao et al. (2025) and Jamal et al. (2021), by demonstrating that ensemble techniques can effectively mitigate the “black box” problem when paired with rigorous explainability frameworks.

Beyond predictive metrics, the integration of SHAP analysis provided critical insights into the features driving these predictions, bridging the gap between model accuracy and interpretability. At the global level, the analysis highlighted that variables such as ethnicity, airbag deployment, the type of first harmful event, and restraint usage are the primary determinants of injury severity across the dataset. The SHAP summary plots elucidated the directionality of these relationships; for instance, the absence of restraint usage and specific harmful events (like collisions with fixed objects) were consistently associated with higher SHAP values, pushing predictions toward fatal outcomes.

This corroborates previous research by Chen et al. (2016) and Ehsani et al. (2023), which identified reckless behaviors and lack of safety equipment as major risk factors. Furthermore, the high importance ranking of ethnicity in our models warrants close attention, as it may serve as a proxy for socio-economic disparities in road infrastructure quality or post-crash medical access, a concern raised by Giron et al. (2025) regarding healthcare outcomes in lower-income communities. By quantifying the magnitude of these contributors, this study confirms that ensemble models can capture the intricate dependencies between driver demographics, vehicle safety features, and environmental conditions that define crash severity.

A unique and critical contribution of this research is the operationalization of local interpretability, addressing a significant limitation in existing traffic safety studies that predominantly focus on global trends. Using force plots and waterfall plots, we demonstrated the ability to dissect individual crash scenarios, revealing how specific feature values, such as a driver’s age or a specific roadway functional classification, interact to produce a specific prediction for a single event. For example, the visual analysis of specific samples showed how protective factors, like airbag deployment, could “pull” a prediction down to a less severe category, while risk factors like alcohol involvement “pushed” it toward fatality. This level of granularity is essential for transforming machine learning outputs into actionable tools for first responders and transportation planners. Unlike global metrics which inform broad policy, these local explanations allow stakeholders to understand the “why” behind a specific high-risk prediction in real-time settings, fulfilling the need for context-aware decision support systems highlighted by Merabet et al. (2025a).

Despite these promising results, this study is subject to certain limitations that offer avenues for future research. The models were developed using data from the National Highway Traffic Safety Administration (NHTSA), and while the dataset is extensive, it is susceptible to the inherent inconsistencies, missing values, and reporting errors common in large-scale police-reported crash databases. Furthermore, the study focused on a specific five-year period (2018–2022) which included anomalous traffic patterns caused by the COVID-19 pandemic; while the models performed well, the long-term generalizability of these specific feature weights across different post-pandemic traffic behaviors remains to be fully tested. Additionally, while ensemble models like HistGBRT and XGBoost offer superior accuracy, their computational complexity is higher than simpler statistical methods, which may present challenges for deployment on resource-constrained edge devices used in real-time traffic management.

Future work should aim to extend this framework by integrating real-time, dynamic data sources such as weather feeds, traffic flow sensors, and telematics to enhance the temporal resolution of predictions. There is also significant potential in exploring hybrid deep learning architectures that combined with XAI methods could further refine predictive accuracy for non-fatal injury classes where some misclassification persisted. Finally, given the strong predictive influence of demographic factors observed in the global SHAP analysis, future studies should employ causal inference techniques to disentangle the root causes of these disparities, whether they stem from infrastructure inequities, vehicle discrepancies, or systemic issues, to better inform equitable traffic safety policies.

## 5. Conclusion

This study successfully introduced a comprehensive framework for predicting traffic accident injury severity by leveraging high-performance ensemble ML algorithms and explainable AI (XAI) techniques. Through the rigorous evaluation of models such as HistGBRT, XGBoost, and CatBoost, we demonstrated that ensemble architectures consistently achieve superior predictive reliability, specifically attaining a perfect 100% sensitivity in identifying fatal injuries, a critical requirement for safety-sensitive domains.

Our work’s primary contribution lies in the systematic integration of SHAP to achieve dual-level interpretability, effectively transforming “black-box” models into transparent decision-support tools. By utilizing global SHAP summary plots, we identified that features such as ethnicity, airbag deployment status, and the type of first harmful event are the most influential determinants of crash outcomes across the population. Simultaneously, the application of local SHAP waterfall and force plots provided instance-level rationales, allowing for a granular understanding of how specific risk factors—such as seatbelt non-compliance or road characteristics—contributed to the severity of an individual accident.

These findings offer distinct implications for different stakeholders. For policymakers and transportation planners, the global feature importance rankings provide a data-driven foundation for prioritizing safety interventions, such as improving restraint usage or addressing disparities in road infrastructure highlighted by demographic variables. For first responders and emergency analysts, the local interpretability framework offers real-time insights into the specific drivers of injury for a given incident, potentially facilitating faster and more informed medical and operational decisions. Furthermore, the temporal analysis of NHTSA data between 2018 and 2022 revealed a concerning 41.5% increase in fatal injuries post-pandemic, reinforcing the urgency of deploying such interpretable predictive tools to mitigate the rising trend of road fatalities.

In summary, this research bridges the critical gap between high-accuracy machine learning performance and the transparency required for operational safety applications. While the study is limited by the constraints of police-reported data and the computational complexity of ensemble methods, it provides a generalizable framework for the deployment of interpretable ML tools in safety-critical domains. Future work should focus on integrating real-time telematics and environmental data to further enhance the temporal resolution and prescriptive value of these models, ultimately supporting global efforts to reduce traffic-related injuries and deaths.

## Author Contributions

E.Z.: conceptualization, methodology, software, data curation, writing—original draft preparation and visualization; O.M.: conceptualization, methodology, validation, data curation, visualization, formal analysis, and writing—original draft preparation; I.D.: writing—review and editing, validation, supervision, project administration, and funding acquisition. All authors have read and agreed to the published version of the manuscript.

## Data Availability Statement

The data used in this study are available from the US National Highway Traffic Safety Administration (NHTSA) Fatality Analysis Reporting System (FARS) at https://www.nhtsa.gov/file-downloads?p=nhtsa/downloads/FARS/ (accessed on 15 January 2024).

